# No evidence for a causal link between plasma lipids and multiple sclerosis susceptibility and severity: a Mendelian randomisation study

**DOI:** 10.1101/2025.10.27.25338893

**Authors:** Jakub Nagrodzki, Petroula Proitsi, Manuela Tan, Ruth Dobson, Benjamin M Jacobs

**Author notes:** **Correspondence to:** Dr Benjamin Jacobs, Centre for Preventive Neurology, Wolfson Institute of Population Health, Queen Mary University of London, UK.

## Abstract

Observational studies suggest that alterations in circulating lipid levels may be associated with multiple sclerosis (MS) susceptibility and severity. Our study employs two-sample Mendelian randomization (MR) to investigate whether these relationships are causal.

Genetic instruments for 249 metabolites, predominantly lipids and lipoproteins, were obtained from a combined dataset of European-ancestry individuals from the UK and Estonian Biobanks. Outcome data were obtained from the International MS Genetics Consortium GWAS studies of MS susceptibility and severity. MR analyses considered MS susceptibility and severity as distinct outcomes and utilized the inverse variance-weighted multiplicative random-effects method in the main analysis.

No metabolic exposures demonstrated statistically significant evidence of a causal relationship with MS susceptibility. For MS severity, two traits showed suggestive associations – triglycerides in very-low-density lipoproteins (VLDL) (β = -0.101, *SE* = 0.028, *p* = 3.9 × 10^-4^) and triglycerides in chylomicrons and extremely large VLDL (β = -0.104, *SE* = 0.032, *p* = 1.2 × 10^-3^). The MR-Egger intercept suggested that these observations may be driven by horizontal pleiotropy. Sensitivity analysis with multivariable MR using Body Mass Index as a possible confounder demonstrated substantial attenuation of instrument strength once genetic overlap with BMI was taken into account.

In this study we found no convincing evidence that circulating lipids or lipid-related metabolites exert a causal influence on susceptibility to, or severity of, multiple sclerosis. The nominally statistically significant result is likely a result of horizontal pleiotropy.

## Introduction

Observational evidence suggests an association between alterations in circulating lipids and lipoproteins and the clinical course of multiple sclerosis (MS).^1^ Several established MS susceptibility genes are thought to exert their role through altering lipid metabolism.^2,3^ For example, 7-dehydrocholesterol reductase encoded by the *DHCR7* gene, a key enzyme in cholesterol biosynthesis,^4^ has been reported as an MS susceptibility locus.^5^

The progression of disability in MS may also be linked to plasma lipids and lipoproteins. Higher plasma concentrations of low-density lipoprotein cholesterol, lipids within the very-low density lipoprotein (VLDL) subclasses, total cholesterol, apolipoprotein B, and triglycerides correlate with inflammatory MRI measures or greater disability on the Expanded Disability Status Scale (EDSS).^6,7^ However, trials which attempted to use medication altering lipid profiles, such as statins, to change the natural history of the disease have so far been negative.^8,9^

It is plausible that the observed changes in the levels of plasma lipids in MS may be a consequence of the disease process, cardiometabolic sequelae of disability, or be related to treatments (such as interferon-β,^10^ fingolimod,^11^ or even corticosteroids^12^) rather than be causally involved in MS susceptibility and disability progression.

Mendelian randomisation (MR) uses inherited genetic variants that influence a risk factor (e.g., lipid levels) as natural experiments to test whether changes in that factor cause differences in disease. Because these variants are fixed at conception and largely independent of lifestyle or disease processes, MR is less prone to confounding and reverse causation, making it well-suited to test whether plasma lipids contribute causally to MS pathogenesis and progression, rather than simply reflecting the disease or its treatments.

Here, we apply two-sample MR to large-scale genome-wide association data to test whether genetically elevated plasma lipids and lipoproteins influence the risk of developing MS or the subsequent trajectory of disability.

## Methods

### Outcome GWAS

Summary statistics for MS susceptibility and severity were obtained from published International Multiple Sclerosis Genetics Consortium GWAS studies.^13,14^ The discovery-stage susceptibility GWAS reported autosomal SNPs with minor allele frequency (MAF) > 0.01 and imputation quality > 0.1 from 14,802 subjects with MS and 26,703 controls.^14^ The severity GWAS correlates autosomal SNPs with MAF > 0.01 from 12,584 people with MS to age-related multiple sclerosis severity score (ARMSS).^13^ Both datasets were available in the hg19 genome build, were converted to hg38, and comprised participants of European ancestry.

### Exposure GWAS

Metabolite-level genetic associations were obtained from a metabolome genome-wide association study (GWAS), which combined 599,249 participants of European ancestry from the UK Biobank and the Estonian Biobank (meta_EUR).^15^ The study encompassed 249 circulating metabolites, out of which 232 corresponded to lipid-related metabolites (fatty acids, cholesterol, ketone bodies, lipids or lipoproteins). All metabolites were considered, without an *a priori* focus. The full list of metabolites and their categorisation is provided in supplementary table S5 and is in line with that used in the original paper.^15^

Instruments were restricted to autosomal, biallelic single-nucleotide polymorphisms (SNPs) with MAF > 0.05, imputation quality > 0.9 and reaching a genome-wide significance of *p* < 5 × 10□□ for association with the exposure. Palindromic variants were excluded to prevent strand ambiguity. Instruments were then matched with those present in the outcome dataset and only instruments present in both the outcome and the exposure datasets after filtering were included for analysis.

### Harmonisation and instrument selection

Harmonisation was performed by aligning exposure and outcome datasets on chromosome, position, reference and alternative alleles to ensure that effect estimates corresponded to the same allele in both datasets. Linkage-disequilibrium (LD) clumping was carried out with PLINK using the 503 European-ancestry participants present in the 1000 Genomes Project reference panel,^16^ lifted to hg38 coordinates using LiftOver, as the LD reference. We retained the index variant with the smallest *p* value in each 10 Mb window when pairwise *r*^*2*^ exceeded 0.001. The resulting independent SNPs formed the genetic instruments for every metabolite (final SNPs number reported in Supplementary tables S1 and S2; all the instruments used in the main analysis of MS severity reported in Supplementary table S6).

### Mendelian randomisation analyses

Two-sample Mendelian randomisation was performed with the TwoSampleMR R package^17^, in line with current consensus guidelines.^18^ In brief, for each metabolite, the primary causal estimate was derived with the inverse-variance-weighted model under a multiplicative random-effects assumption. Cochran’s Q statistic was used to quantify heterogeneity. Horizontal pleiotropy was assessed by the MR-Egger intercept. MR-PRESSO was used for further sensitivity analysis as a pleiotropy-robust method. Metabolites suggestive of statistically significant results also underwent the leave-one-out analysis. All analyses were repeated separately for MS susceptibility and MS severity.

### Correction for multiple comparisons

In order for the correction for multiple comparisons to take into account the correlations between circulating metabolite levels, we calculated the effective number of independent tests (Meff). The metabolite-metabolite correlation matrix reported by Tambets et al.^15^ was processed with the eigenvalue method implemented in the *poolr* R package.^19,20^ The resulting effective number of tests (33) served as the denominator in a Bonferroni correction.

Statistical significance was declared at two thresholds: □_*Meff*_ < 1.5 × 10^-3^ (0.05 / 33) and full Bonferroni correction □_*Bonf*_ < 2 × 10^-4^ (0.05 / 249).

### Instrument strength and power

For every instrument we quantified the proportion of exposure variance explained (*R*^*2*^) and the corresponding first-stage F-statistic. *R*^*2*^ for each SNP was computed as β^2^ × 2 × EAF × (1 − EAF), where β is the SNP-metabolite effect estimate and EAF is the effect-allele frequency. Metabolite-specific *R*^*2*^ values were summed to obtain the total *R*^*2*^ per instrument. The F-statistic was calculated as (R^2^ × [N − k − 1]) / [k × (1 − R^2^)], where *N* denotes the exposure GWAS sample size and *k* the number of SNPs. Instruments with *F* > 10 were considered sufficiently strong.

### Sensitivity analyses

When the univariable analyses suggested a potentially causal association, we conducted multivariable Mendelian randomisation (MVMR) to assess whether the signal could be explained by horizontal pleiotropy operating through correlated traits. For each metabolite, we first identified a plausible correlate that might confound the SNP-outcome relation; the rationale and data sources are detailed in the Results section. Summary statistics for all variables were filtered with the same criteria described for the univariable analysis, after which the harmonised SNP list was merged across exposures and outcomes. MVMR was carried out in R with the *mvmr* package.^21^

Instrument strength in the multivariable setting was evaluated with the conditional *F* statistic, which extends the ordinary first-stage *F* statistic to multiple exposures by testing the association of each set of SNP coefficients with its target exposure, conditional on the remaining exposures.^22^ Conditional *F* statistic values below 10 were taken to indicate weak instruments.

As a further control, we used the Global Lipids Genetics Consortium (GLGC) GWAS^23^ of five lipid traits to extract genetic instruments and conducted a two-sample MR analysis, using the same methodology as described above.

### Data availability and transparency

Exposure summary statistics are available from the GWAS Catalog (https://www.ebi.ac.uk/gwas/, accession numbers GCST90451106 - GCST90451354). Outcome summary statistics were provided by the IMGSC and can be applied for online (https://imsgc.net). Analysis code is available at https://github.com/jaknag/ms_lipidomics.

## Results

### MS susceptibility

No metabolite met the corrected significance threshold in the primary MR analysis of MS susceptibility (Fig. 1A). All instruments were sufficiently strong (*F* statistics 56.8 - 267). Full results are available in Supplementary table S1. A control analysis using five lipid classes from the GLGC GWAS likewise detected no associations with MS susceptibility (all *p* values > 0.3, Supplementary table S3).

**Fig 1.**
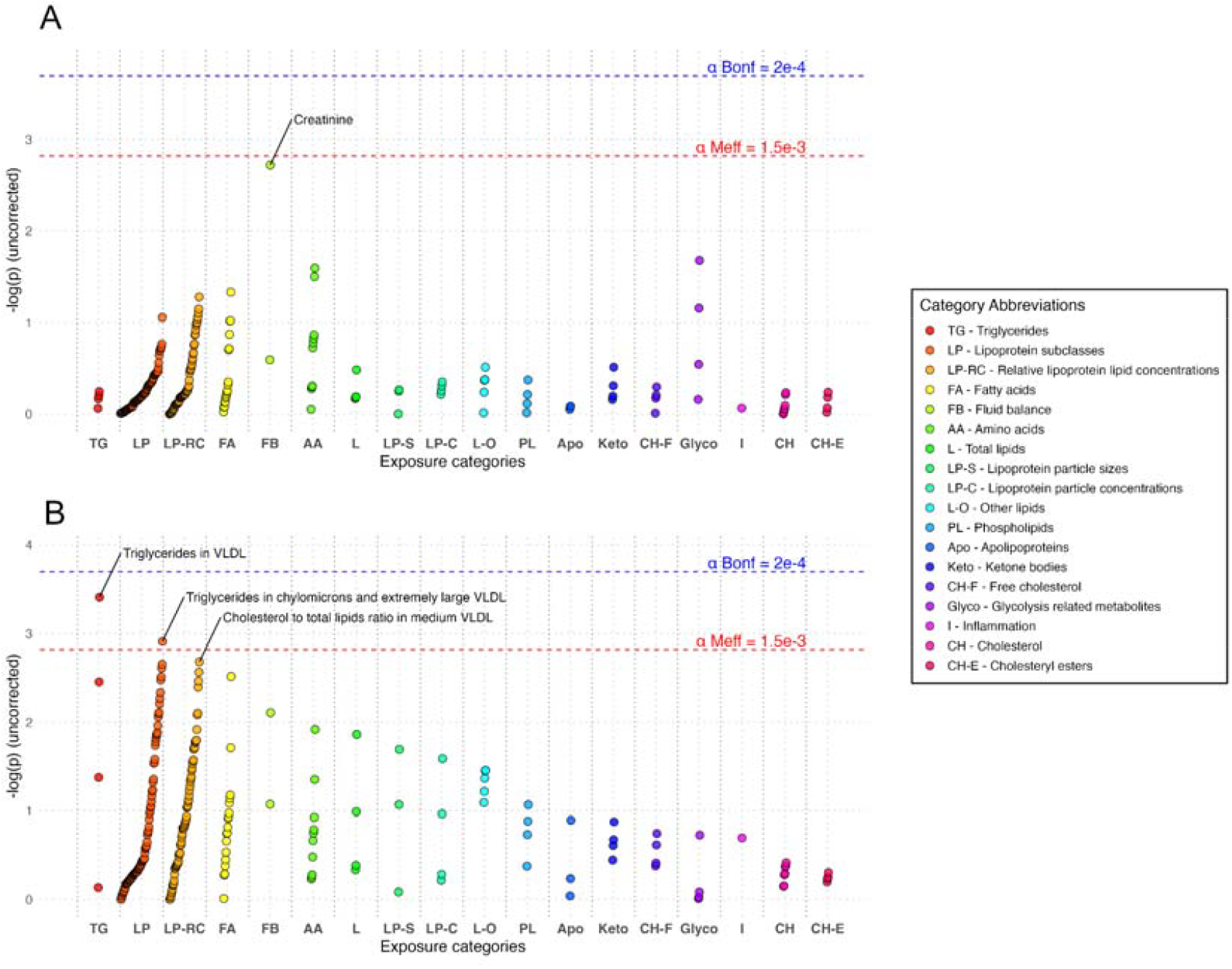
A Manhattan-style plot showing the associations between exposures and (A) MS susceptibility or (B) MS severity. Exposures are grouped by categories, according to their classification in the original paper,^15^ also included in Supplementary Table 5. Horizontal lines indicate *p* value thresholds: blue line □_*Bonf*_ = 2 × 10^-4^, and red line □_*Meff*_ = 1.5 × 10^-3^.

### MS severity

In the analysis of MS severity, two triglyceride-based exposures crossed the □_*Meff*_ threshold (Fig. 1B). Genetically predicted triglycerides in very-low-density lipoproteins (VLDL_TG) were inversely correlated with MS severity (β = -0.101, *SE* = 0.028, *p* = 3.9 × 10^-4^). These results suggest that a genetically-predicted increase in plasma VLDL_TG by 1 standard deviation was associated with a 0.1 point reduction in ARMSS, corresponding to a reduction of 0.03 EDSS points at age 40 and EDSS 3.0 (a small effect magnitude given that the minimum EDSS step is 0.5). A similar effect was observed for triglycerides in chylomicrons and extremely large VLDL (XXL_VLDL_TG; β = -0.104, *SE* = 0.032, *p* = 1.2 × 10^-3^). Neither of these were significant at the full Bonferroni corrected threshold □_*Bonf*_.

Instrument heterogeneity was not detected for either exposure (Cochran’s Q, *p*_*Q*_s > 0.070). The MR-Egger intercept suggested possible directional pleiotropy for VLDL_TG (intercept = -0.002, *p* = 0.066) and for XXL_VLDL_TG (intercept = -0.004, *p* = 0.001). Although the pleiotropy-robust method MR-PRESSO suggested significant results (both *p* values < 1.34 × 10^-3^), post-hoc analyses with MR Egger, weighted median, and weighted mode methods indicated inconsistent direction of effect size, and were not significant for both VLDL_TG (MR Egger β = -0.035, *p* = 0.439; weighted median β = -0.128, *p* = 7.54 × 10^-3^; weighted mode β = -0.113, *p* = 0.018) and XXL_VLDL_TG (MR Egger β = 0.033, *p* = 0.526; weighted median β = -0.112, *p* = 0.041; weighted mode β = -0.082, *p* = 0.171).

Instrument strength remained robust across all 249 metabolites (*F* 55.8 - 311.8), and leave-one-out analyses revealed no single variant driving the associations. Full results are available in Supplementary table S2.

Because body-mass index (BMI) has been linked to both VLDL_TG concentrations and clinical course in multiple sclerosis,^24^ we conducted multivariable MR to determine whether the putative circulating metabolite effect on severity was confounded by BMI. SNP-BMI associations from a large European-ancestry BMI GWAS^25^ were analysed jointly with each lipid trait and MS severity. In the model which included VLDL_TG, the conditional *F* statistic fell well below the conventional threshold of 10 for both exposures (*F*_*BMI*_ = 6.49; *F*_*VLDL_TG*_ = 3.82), signalling substantial attenuation of instrument strength once genetic overlap with BMI was taken into account. Although the multivariable regression yielded a nominally protective coefficient for VLDL_TG (β = -0.026, *SE* = 0.0021, *p* = 1.4 × 10□^3^□), the weak conditional *F* statistic renders this estimate unreliable. The same pattern was observed when XXL_VLDL_TG were used as the lipid exposure – conditional *F* statistics again indicated weak instruments (*F*_*BMI*_ = 6.39; *F*_*XXL_VLDL_TG*_ = 3.07).

A replication analysis using genetic instruments for five lipid classes, including triglycerides, from the GLGC GWAS showed no associations with MS severity (all *p* values > 0.5 uncorr., Supplementary table S4).

## Discussion

In summary, in this two-sample Mendelian randomisation study we found no convincing evidence that circulating lipids or lipid-related metabolites exert a causal influence on the severity of, or susceptibility to, multiple sclerosis.

Although triglycerides in VLDL and triglycerides in chylomicrons and extremely large VLDL passed the □_*Meff*_ statistical threshold corrected for the number of effective tests in the severity analysis, these signals likely represent false positives for several reasons. First, the apparent negative correlation between triglyceride content in VLDL and chylomicrons and MS severity is contrary to evidence linking VLDL lipids to heightened neuroinflammation and demyelination.^26^ Second, the MR-Egger intercept for both exposures was different from zero, signalling horizontal pleiotropy, which can distort causal estimates in an MR analysis, especially when inverse-variance method is used.^18^ This consideration is especially relevant given the pervasive pleiotropy in the genetic architecture of lipid traits. Third, post-hoc sensitivity analyses with outlier-robust methods did not suggest any significant effects. Fourth, the multivariable analysis demonstrates that effect size estimates are significantly attenuated by inclusion of BMI, which suggests that the main effect may be, at least partly, attributable to residual confounding by BMI or other pleiotropic pathways, however interpretation of these results is challenging due to the low conditional *F* statistic. Finally, these results did not surpass a stringent Bonferroni correction □_*Bonf*_.

Several limitations exist which limit the interpretation of these findings. First, the likely effects – if they do exist – are small and existing GWAS studies may not be sufficiently powered to detect them or results may depend on the granularity of datasets used.^27^ Second, we interrogated metabolites in peripheral blood, yet this is a distal compartment in Multiple Sclerosis, and it is plausible that if dysregulation of circulating lipids does exert a causal effect on MS risk or severity, these effects might be more readily discovered by studying cerebrospinal fluid (CSF) or brain metabolomics. Lipid exchange across the blood-brain barrier is tightly regulated, and CSF lipid profiles differ appreciably from those in serum.^28^ Third, existing studies provide a snapshot of exposure but cannot capture the short-term diurnal and post-prandial fluctuations that characterise lipid metabolism.^29,30^ Finally, both the exposure and the outcome are complex, polygenic traits embedded in dense correlation networks. Horizontal pleiotropy is therefore pervasive, and stringent multiple-comparison control invariably trades statistical power for specificity. Our use of the effective-test procedure could not fully guard against type I error, as illustrated by the probable false positive signals detected, while more conservative thresholds likely risk substantial type II error.

Overall, our large-scale two-sample MR study contributes to the growing body of evidence that circulating lipids and lipoproteins are unlikely to be causally involved in MS susceptibility or severity, and certainly argues against the presence of an obvious ‘smoking gun’ lipid mediator which could be straightforwardly targeted. Future work could include: assessment of CSF lipid and lipoprotein profiles rather than plasma, capturing dynamic lipid states with diurnal and post-prandial assessments rather than single static measures, implementing longitudinal sampling to relate changes in lipid profiles to disease activity, and applying multivariable MR to better isolate lipid effects from confounders.

## References

1. Zhornitsky S, McKay KA, Metz LM, et al. Cholesterol and markers of cholesterol turnover in multiple sclerosis: relationship with disease outcomes. Mult Scler Relat Disord 2016; 5: 53–65.

2. International Multiple Sclerosis Genetics Consortium (IMSGC), Beecham AH, Patsopoulos NA, et al. Analysis of immune-related loci identifies 48 new susceptibility variants for multiple sclerosis. Nat Genet 2013; 45: 1353–1360.

3. Yang F, Li X, Wang J, et al. Identification of lipid metabolism-related gene markers and construction of a diagnostic model for multiple sclerosis: An integrated analysis by bioinformatics and machine learning. Anal Biochem 2025; 700: 115781.

4. Prabhu AV, Luu W, Li D, et al. DHCR7: A vital enzyme switch between cholesterol and vitamin D production. Prog Lipid Res 2016; 64: 138–151.

5. Alloza I, Otaegui D, de Lapuente AL, et al. ANKRD55 and DHCR7 are novel multiple sclerosis risk loci. Genes Immun 2012; 13: 253–257.

6. Weinstock-Guttman B, Zivadinov R, Horakova D, et al. Lipid profiles are associated with lesion formation over 24 months in interferon-β treated patients following the first demyelinating event. J Neurol Neurosurg Psychiatry 2013; 84: 1186–1191.

7. Tettey P, Simpson S, Taylor B, et al. An adverse lipid profile is associated with disability and progression in disability, in people with MS. Mult Scler Houndmills Basingstoke Engl 2014; 20: 1737–1744.

8. Almramhi MM, Finan C, Storm CS, et al. Exploring the Role of Plasma Lipids and Statin Interventions on Multiple Sclerosis Risk and Severity. Neurology 2023; 101: e1729– e1740.

9. Blackstone J, Williams T, Nicholas JM, et al. Evaluating the effectiveness of simvastatin in slowing the progression of disability in secondary progressive multiple sclerosis (MS-STAT2): protocol for a multicentre, randomised controlled, double-blind, phase 3 clinical trial in the UK. BMJ Open 2024; 14: e086414.

10. Uher T, Fellows K, Horakova D, et al. Serum lipid profile changes predict neurodegeneration in interferon-β1a-treated multiple sclerosis patients. J Lipid Res 2017; 58: 403–411.

11. Rauma I, Huhtala H, Soilu-Hänninen M, et al. Lipid Profile Alterations during Fingolimod Treatment in Multiple Sclerosis Patients. J Neuroimmune Pharmacol Off J Soc NeuroImmune Pharmacol 2020; 15: 567–569.

12. Choi HK, Seeger JD. Glucocorticoid use and serum lipid levels in US adults: The third national health and nutrition examination survey. Arthritis Care Res 2005; 53: 528–535.

13. Harroud A, Stridh P, McCauley JL, et al. Locus for severity implicates CNS resilience in progression of multiple sclerosis. Nature 2023; 619: 323–331.

14. International Multiple Sclerosis Genetics Consortium. Multiple sclerosis genomic map implicates peripheral immune cells and microglia in susceptibility. Science 2019; 365: eaav7188.

15. Tambets R, Kronberg J, Graaf A van der, et al. Genome-wide association study for circulating metabolic traits in 619,372 individuals. 2025; 2024.10.15.24315557.

16. Byrska-Bishop M, Evani US, Zhao X, et al. High-coverage whole-genome sequencing of the expanded 1000 Genomes Project cohort including 602 trios. Cell 2022; 185: 3426-3440.e19.

17. Hemani G, Zheng J, Elsworth B, et al. The MR-Base platform supports systematic causal inference across the human phenome. eLife 2018; 7: e34408.

18. Burgess S, Davey Smith G, Davies NM, et al. Guidelines for performing Mendelian randomization investigations: update for summer 2023. Wellcome Open Res 2019; 4: 186.

19. Cinar O, Viechtbauer W. The poolr Package for Combining Independent and Dependent p Values. J Stat Softw 2022; 101: 1–42.

20. Li J, Ji L. Adjusting multiple testing in multilocus analyses using the eigenvalues of a correlation matrix. Heredity 2005; 95: 221–227.

21. Sanderson E, Davey Smith G, Windmeijer F, et al. An examination of multivariable Mendelian randomization in the single-sample and two-sample summary data settings. Int J Epidemiol 2019; 48: 713–727.

22. Sanderson E, Spiller W, Bowden J. Testing and correcting for weak and pleiotropic instruments in two-sample multivariable Mendelian randomization. Stat Med 2021; 40: 5434–5452.

23. Graham SE, Clarke SL, Wu K-HH, et al. The power of genetic diversity in genome-wide association studies of lipids. Nature 2021; 600: 675–679.

24. Jorissen W, Wouters E, Bogie JF, et al. Relapsing-remitting multiple sclerosis patients display an altered lipoprotein profile with dysfunctional HDL. Sci Rep; 7. Epub ahead of print 23 February 2017. DOI: 10.1038/srep43410.

25. Verma A, Huffman JE, Rodriguez A, et al. Diversity and scale: Genetic architecture of 2068 traits in the VA Million Veteran Program. Science 2024; 385: eadj1182.

26. Gafson AR, Thorne T, McKechnie CIJ, et al. Lipoprotein markers associated with disability from multiple sclerosis. Sci Rep 2018; 8: 17026.

27. Gilchrist L, Mutz J, Hysi P, et al. Evaluating metabolome-wide causal effects on risk for psychiatric and neurodegenerative disorders. BMC Med 2025; 23: 326.

28. Tsujita M, Melchior JT, Yokoyama S. Lipoprotein Particles in Cerebrospinal Fluid. Arterioscler Thromb Vasc Biol 2024; 44: 1042–1052.

29. Dallmann R, Viola AU, Tarokh L, et al. The human circadian metabolome. Proc Natl Acad Sci U S A 2012; 109: 2625–2629.

30. Yan S, Li L, Horner D, et al. Characterizing human postprandial metabolic response using multiway data analysis. Metabolomics Off J Metabolomic Soc 2024; 20: 50.

